# Data presented by the UK government as lockdown was eased shows the transmission of COVID-19 had already increased

**DOI:** 10.1101/2020.06.28.20141960

**Authors:** Mike Lonergan

## Abstract

**Background:** Coronavirus disease 2019 (COVID-19) is an international emergency that has been addressed in many countries by changes in and restrictions on behaviour. These are often collectively labelled social distancing and lockdown. On the 23^rd^ June 2020, Boris Johnson, the Prime Minister of the United Kingdom announced substantial easings of restrictions. This paper examines some of the data he presented.

**Methods:** Generalised additive models, with negative binomial errors and cyclic term representing day-of-week effects, were fitted to data on the daily numbers of new confirmed cases of COVID-19. Exponential rates for the epidemic were estimated for different periods, and then used to calculate R, the reproduction number, for the disease in different periods.

**Results:** After an initial stabilisation, the lockdown reduced R to around 0.81 (95% CI: 0.79, 0.82). This value increased to around 0.94 (95% CI 0.89, 0.996) for the fortnight from the 9^th^ June 2020.

**Conclusions:** Official UK data, presented as the easing of the lockdown was announced, shows that R was already more than half way back to 1 at that point. That suggests there was little scope for the announced changes to be implemented without restarting the spread of the disease.

## Introduction

Like most other communicable diseases, coronavirus disease 2019 (COVID-19) is primarily a public health issue. The medical care of infected individuals is, of course, very important but most of those who become infected with SARS-CoV-2[1] neither receive nor need such care. The major determinant of mortality and morbidity is the number of individuals exposed to infection.

Changes in and restrictions on public behaviour, which have often been collectively labelled social distancing and lockdown, were adopted in most countries as they have attempted to reduce the impact of COVID-19. While the details have varied, the overall effects have been quite similar in countries that managed to alter the course of the epidemic [2] with R, the reproduction number, for the disease falling from around 3 to a little below 1.

R is the number of individuals that one infected individual can be expected to pass the disease to, over the course of their infectious period. It is critical to disease spread: above 1 an epidemic will accelerate, below that the outbreak will fade away. Its initial value, when everyone in a population is susceptible to infection, is referred to as R_0_. If a proportion of a population develops immunity, either through natural exposure to the disease or vaccination against it, then R_t_, the effective value of R at that time, will also decrease by that proportion. Despite the large absolute numbers of infections and deaths from COVID-19, only small proportions of national populations have yet been infected so such immunity can be ignored in simple examinations of short-term patterns of infection.

The entry of the United Kingdom into lockdown was slightly messy and idiosyncratic. The details, and timing, have been discussed many times. The overall effect was a slightly less restrictive lockdown than in many other European countries, but with 2m social distancing rather than the 1m suggested by WHO [3, 4]. On the 23^rd^ June 2020, Boris Johnson, the UK Prime Minister presented a revised set of instructions in a press conference. These reduced the distancing requirements and laid out plans for the opening of many commercial enterprises. During the presentation, the trajectory of the epidemic was shown in various slides. One of these showed the numbers of newly confirmed cases from each day throughout the epidemic (Figure 1). At the right hand side of the graph, the decline in the numbers of cases seems to slow. This paper investigates whether that apparent change is real, and what it may indicate.

**Figure 1:**
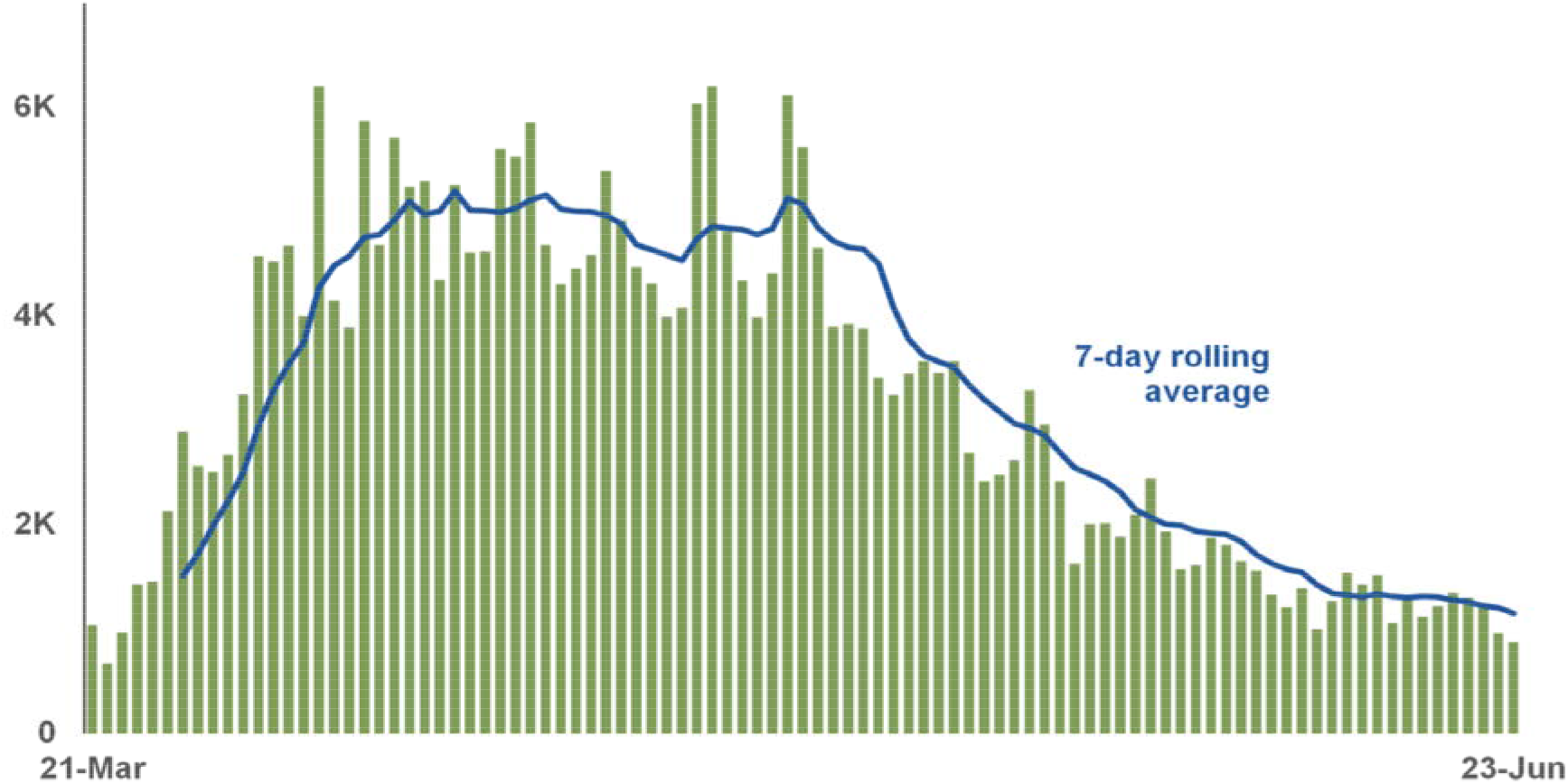
The number of newly confirmed cases of COVID-19 for each day, as shown at the UK Governmental briefing on 23 June 2020. Taken from https://assets.publishing.service.gov.uk/government/uploads/system/uploads/attachment_data/file/894594/2020-06-23_COVID-19_Press_Conference_Slides_with_Annex.pdf

## Methods

The data were downloaded from the government website (https://www.gov.uk/government/publications/slides-and-datasets-to-accompany-coronavirus-press-conference-23-june-2020). Generalised additive models [5] (a form of regression model that allows for curves as well as straight lines) were fitted to the data with negative binomial errors and a cyclic smooth to capture day-of-week patterns. The analysis was carried out in R 4.0.0 [6], and the R script is available as a supplement to the online version of this article. The day-of-week effect was included as a nuisance that needed to be dealt with, rather than being of any intrinsic interest here, and so is largely ignored in the remained of the paper. All the model predictions treat every day as a Wednesday.

A model was fitted to all the data with a 20-knot thin-plate spline for date (Figure 2). It seemed to show a change around June 9^th^, so the model was refitted to just the data from before then. The divergence of the predictions of the two models after June 9^th^ suggests that there is a real difference. Examination of the model trajectory suggested that could be divided into four periods, and exponential patterns fitted to each one. The exponential model depends on two simplifications: that behaviour affecting disease transmission remains constant within each period, and that the proportion of the population still susceptible to infection remains constant. This second simplification is based on only a small proportion of the total population being infected in each period, which is plausible for all except the second period. Estimates, and 95% confidence intervals, were extracted for the slope in each of the periods, and converted into estimates of R. The second period, starting from around the date of lockdown, contained dramatic changes in the UK’s testing strategies and is very difficult to interpret.

**Figure 2:**
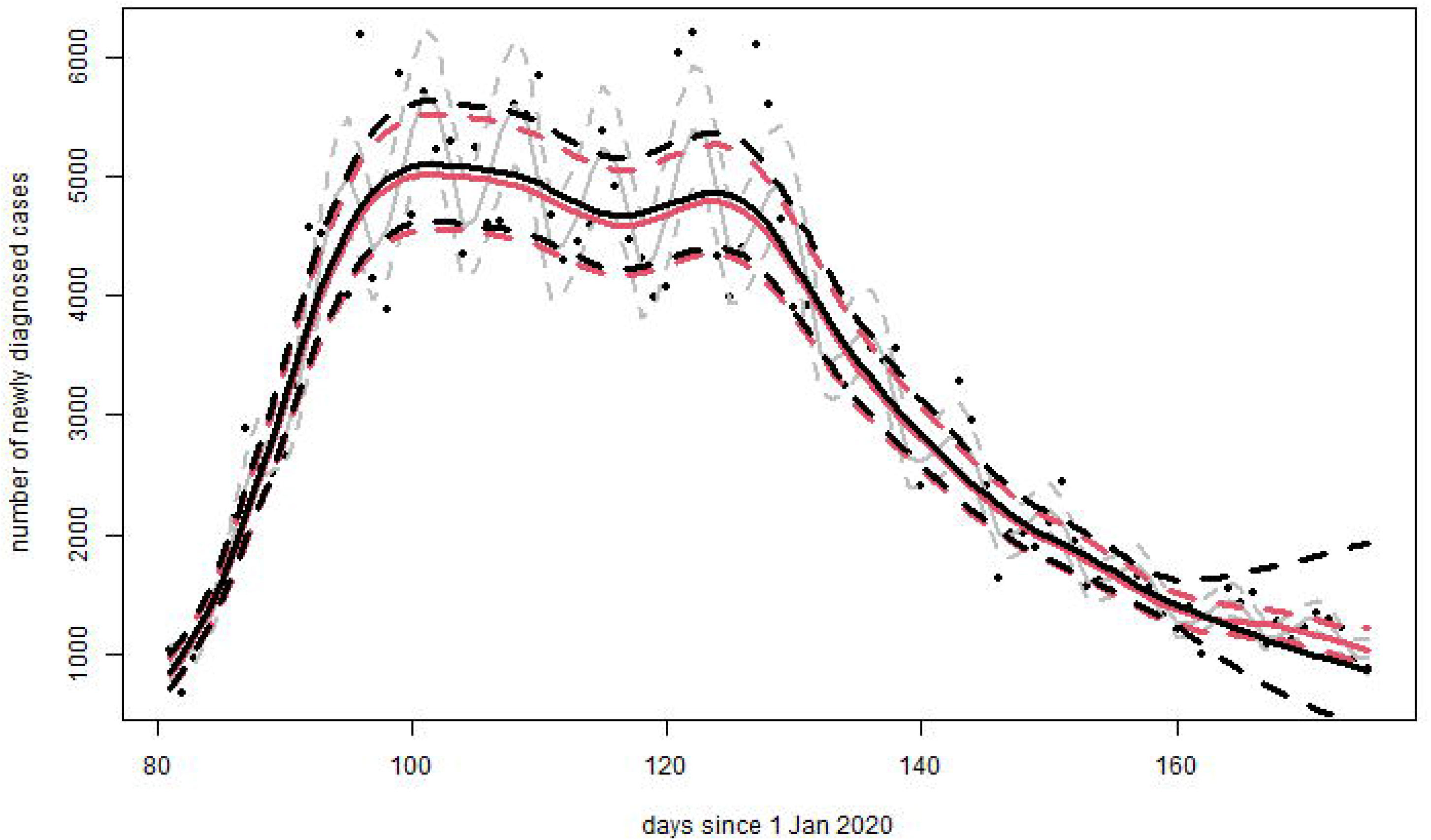
generalised additive models of trajectory in COVID-19 cases in the UK. The grey lines show the mean and 95% confidence intervals estimated for each day by the model fitted to all the data and including the day-of week effect. The other lines predict as if every day were a Wednesday. The black lines are for predictions using the data up to 9 June 2020. The red lines are from fitting the model to the complete dataset.

Because these data include only a fraction of the cases in the country, they cannot be directly used to estimate R. Instead, the R0 library[7] was used to apply the method of Wallinga and Lipsitch[8] to convert the estimates, and associated uncertainties, of the exponential trajectories into estimates of R. This approach requires an estimated distribution for the serial interval of infection. The lognormal with mean 4 days and standard deviation of 2.9 days calculated by Nishiura, Linton and Akhmetzhanov[9] was used for this. While changing this distribution changes the individual values estimated for R, the relationship between the estimates is relatively insensitive to plausible choices.

## Results

Figures 2 and 3 show the patterns of cases estimated by the models. The smooth model of the full dataset captures 96% of the deviance, while the exponential model captures 95%.The lower bound of the 95% confidence interval for values predicted for dates after June 9^th^ from the model fitted to the full dataset is above the predicted values based on data up to June 9^th^, strongly suggesting a real change occurred then.

**Figure 3:**
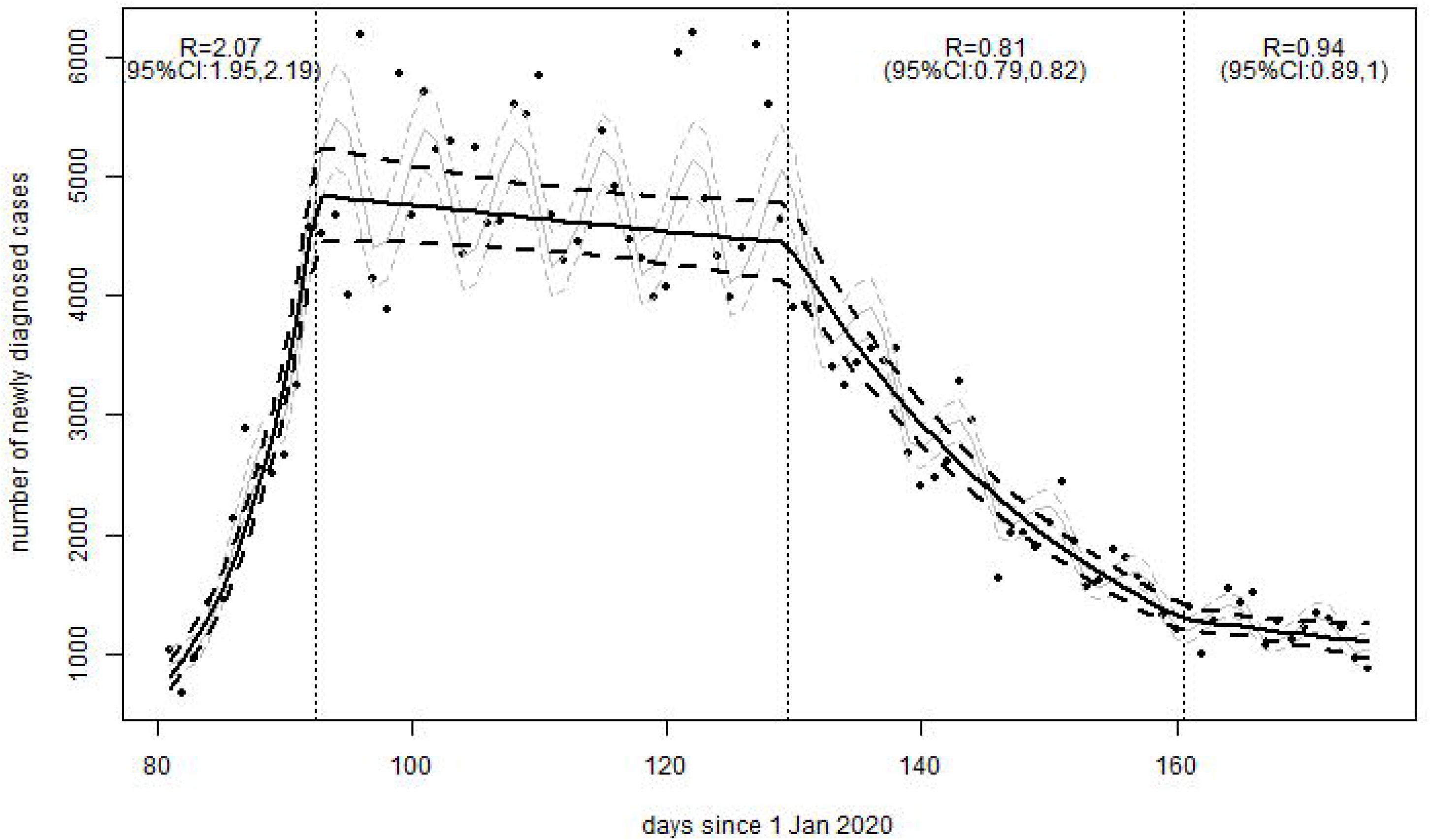
Modelling the outbreak as a series of four exponential periods. The grey lines show the mean and 95% confidence intervals estimated for each day by the model fitted to all the data and including the day-of week effect. The black lines show the exponential pattern, treating every day as a Wednesday. Values of R, the reproduction number for COVID-19, are shown in each period of interest. These, and their confidence intervals, are calculated from the relevant estimates of slopes.

The slopes, and the estimates of R, for the four periods are shown in table 1. The estimate for the first period, of 2.07 is among the lower estimates of R_0_ for COVID-19. Unsurprisingly, given the use of very similar methodologies, it matches that in [2]. The estimate, 0.81, for R in the third period is similar for most other estimates, though its confidence interval is much narrower than that provided by most other models. There is a substantial increase in R for the final period, the estimate is quite distinct from that for the third period, and the upper bound of the final confidence interval is very close to 1.

**Table 1:** Slopes and values of R, the reproduction number, for COVID-19 estimated from the exponential model for the four periods.

| Period | Days (from 01/01/2020) | Dates | Slope (95% CI) | R (95% CI) |
| --- | --- | --- | --- | --- |
| 1 | 81 – 92 | 21/03 – 01/04 | 0.155 (0.141, 0.169) | 2.07 (1.95, 2.19) |
| 2 | 93 – 129 | 02/04 – 08/05 | -0.002 (-0.005, 0.001) | 0.99 (0.97, 1.003) |
| 3 | 130 – 160 | 09/05 – 08/06 | -0.040 (-0.043, -0.036) | 0.81 (0.79, 0.82) |
| 4 | 161 - 175 | 09/06 – 23/06 | -0.011 (-0.022, -0.001) | 0.94 (0.89,0.996) |

## Discussion

### Main finding of this study

This study shows that data available to, and being presented by, the UK government as it described its plans for easing the lockdown in the UK showed that R had increased two weeks before the announcement. This increase had absorbed around two-thirds of the leeway between the value of R during the main period of epidemic decline and 1, suggesting that more than half the safe amount of increase in contact between individuals had already occurred. It also provides evidence that there remained very little room for manoeuvre by that point.

### What is already known on this topic

The values of R for COVID-19, both initially and during the main period of lockdown has been calculated in many ways and many countries. In the countries where the spread of the disease was interrupted, the decline in numbers of cases and mortalities was generally much slower than the preceding increase, suggesting that R had not been pushed far below 1 [2]. News reports, and anecdotal evidence, showed mass attendance at demonstrations and crowded beaches in the UK during June 2020.

### What this study adds

This study shows not only that R for COVID-19 in the UK had increased before the announcement of the easing in lockdown, but that the data demonstrating that change was available, and presented by the prime minister, during his announcement.

### Limitations of this study

Confirmed cases are only people who have been tested for SARS-CoV-2. Only a small proportion of people get tested and this proportion changed dramatically, especially during the early stages of the epidemic. Data for hospitalisations and mortality has fewer problems but are lagged relative to information on cases. This pattern is not yet visible in them. The study contains no information on the relative importance of the various changes that were announced. While it shows that most of the restrictions that affected disease transmission were necessary, it cannot determine which of them actually had any real the consequence. Estimates of the the implications of the easing of the 2m rule, in particular, depend on whether the risk of disease transmission is different at 1 and 2m, a matter that currently depends on the interpretation of the available evidence [3, 4]. These analyses also cannot determine whether the particular circumstances in mid-June can be expected to continue.

## Data Availability

The data are available from a uk government website

https://www.gov.uk/government/publications/slides-and-datasets-to-accompany-coronavirus-press-conference-23-june-2020

## Key points

- transmission of COVID-19in the UK increased around 9^th^ June 2020
- R, the reproduction number for the disease increased from about 0.81 to around 0.94 at that point
- this changes shows that half the space for easing lockdown had been used up by then
- data showing this was presented during the UK government’s announcement easing lockdown on 23^rd^ June 2020.

## References

1. Zhu, N., et al., A Novel Coronavirus from Patients with Pneumonia in China, 2019. New England Journal of Medicine, 2020. 382(8):p. 727–733.

2. Lonergan, M. and J.D. Chalmers, Estimates of the ongoing need for social distancing and control measures post-”lockdown” from trajectories of COVID-19 cases and mortality. European Respiratory Journal, 2020:p. 2001483.

3. Chu, D.K., et al., Physical distancing, face masks, and eye protection to prevent person-to-person transmission of SARS-CoV-2 and COVID-19: a systematic review and meta–analysis. The Lancet.

4. Lonergan, M., Even one metre seems generous. A reanalysis of data in: Chu et al. (2020) Physical distancing, face masks, and eye protection to prevent person-to-person transmission of SARS-CoV-2 and COVID-19. Available at: https://www.researchgate.net/publication/342098022_Even_one_metre_seems_generous_A_reanalysis_of_data_in_Chu_et_al_2020_Physical_distancing_face_masks_and_eye_protection_to_prevent_person-to-person_transmission_of_SARS-CoV-2_and_COVID-19/stats, 2020. p. 8.

5. Wood, S., Generalized Additive Models: An Introduction With R (2nd edition). 2017. 391

6. R_Core_Team, R: A Language and Environment for Statistical Computing, 2020, R Foundation for Statistical Computing: Vienna, Austria.

7. Boelle, P.-Y.O., Thomas, R0: Estimation of R0 and Real-Time Reproduction Number from Epidemics, 2015.

8. Wallinga, J. and M. Lipsitch, How generation intervals shape the relationship between growth rates and reproductive numbers. Proceedings of the Royal Society B-Biological Sciences, 2007. 274(1609):p. 599–604.

9. Nishiura, H., N.M. Linton, and A.R. Akhmetzhanov, Serial interval of novel coronavirus (COVID-19) infections. International journal of infectious diseases: IJID: official publication of the International Society for Infectious Diseases, 2020. 93:p. 284–286.

